# A data-driven assessment of early travel restrictions related to the spreading of the novel COVID-19 within mainland China

**DOI:** 10.1101/2020.03.05.20031740

**Authors:** Alberto Aleta, Qitong Hu, Jiachen Ye, Peng Ji, Yamir Moreno

## Abstract

Two months after it was firstly reported, the novel coronavirus disease COVID-19 has already spread worldwide. However, the vast majority of reported infections have occurred in China. To assess the effect of early travel restrictions adopted by the health authorities in China, we have implemented an epidemic metapopulation model that is fed with mobility data corresponding to 2019 and 2020. This allows to compare two radically different scenarios, one with no travel restrictions and another in which mobility is reduced by a travel ban. Our findings indicate that *i)* travel restrictions are an effective measure in the short term, however, *ii)* they are ineffective when it comes to completely eliminate the disease. The latter is due to the impossibility of removing the risk of seeding the disease to other regions. Our study also highlights the importance of developing more realistic models of behavioral changes when a disease outbreak is unfolding.

## I. INTRODUCTION

In Dec. 31st, 2019, Chinese authorities reported an outbreak of a novel coronavirus disease, called COVID-19 by the World Health Organization. Due to the proximity of the Spring Festival, the Chinese Government implemented quarantine in Wuhan, where the outbreak started, as well as in several nearby cities since Jan. 23rd, 2020. As of Feb. 16th the outbreak is still growing and has already infected 68,584 individuals in China of which 1,666 have died [1]. Much is still unknown about the characteristics of this pathogen. For instance, it remains unclear the animal source of this zoonotic disease, being bats and pangolins currently the two most likely sources [2–4]. It has also been proposed that more than half of the cases might have gone undetected by routinely screening passengers, due to the special characteristics of this disease [5], which makes it possible that infected individuals are asymptomatic while infectious.

Several studies predict a much larger number of infections than the actual number reported by the authorities, claiming that only between 10% and 20% of the cases have been detected and reported [6–9]. The reasons for such deviations between models and actual count of cases are diverse, for instance, the fact that the symptoms could be mild and similar to other flue-like diseases for some people, may induce infected individuals not to seek medical care [10]. On the other hand, on Feb. 13th, 14,840 new cases were reported [11], in contrast to 2,022 cases counted during the previous day [12]. The reason was that previously to that day, only those cases that had been laboratory-confirmed were being recognized as so, whereas from that date onwards, also the clinically diagnosed cases are accounted for. Therefore, the model-based prediction of the numbers of infected individuals can plausibly be larger than the official reports.

From a theoretical and computational point of view, there are groups that have proposed new epidemic models to properly account for the special characteristics of this disease [10, 13, 14]. However, our knowledge of the dynamics of the disease is too limited to be constrained to use such sophisticated models. In fact, in some of these works, the models are fitted to reproduce exactly the reported number of infected individuals, which, as noted before, can be counterproductive given that the actual number of infected individuals in the population is surely higher than those detected either by clinical diagnosis or in the laboratory. Lastly, there has also been intense research aimed at computing the probability that the outbreak extends beyond Wuhan to other cities in China, as well as to the rest of the world [6, 7, 15–18]. These works use historical data in order to produce a risk assessment and obtain probabilities that the disease is imported in other populations. Likewise, modeling efforts have been directed towards gauging the effect of Wuhan’s quarantine on the spreading of the epidemic all over the country (which so far has determined to be a delay of 3 days in the arrival of the peak) and worldwide.

Given the unprecedented characteristics of this outbreak, we here adopt a slightly different approach and study a data-driven metapopulation model that makes use of the actual flows of the population to properly measure the early effects of the travel reductions in China. Specifically, we built a basic metapopulation model [19, 20] of 31 regions in China (except HongKong, Macau and Taiwan). Inside each population, we considered that the individuals in the population interact following a homogeneous mixing scheme. This approach is similar to the ones proposed in [7] and [15]. However, in our case, we perform a data-driven simulation with the actual flows of individuals that have taken during the period of study. That is to say, we do not rely on transportation data (which is most of the time given as the maximum flow capacity between subpopulations), but on a real dataset gathered for this occasion (see Materials and Methods). Given that we restrict the analysis to mainland China, this dataset could be better suited for relatively small scale movements. As recently reported, our modeling framework also allows to make an early assessment of the impact that travel restrictions have had on the spreading of the epidemic in mainland China. The results align with previous findings and indicate that travel restrictions do not have a significant impact in containing the expansion of the disease, though reducing the flow of individuals could lead to a delay in the importation of new cases in other subpopulations. Importantly, ours is another study in which taking the mobility patterns into account in as realistic as possible way emerges as a key factor for modeling purposes. In this sense, our conclusions point out that despite the many advances in disease modeling during the last two decades, there still remain many open challenges, most of them related to how to sensibly incorporate human behavioral changes and response to the COVID-19 outbreak.

## II. MODEL

We implement a stochastic SEIR-metapopulation model to simulate the spread of the epidemic across mainland China. The model can be divided in two discrete processes: the disease dynamics governed by a SEIR compartmental model; and the mobility of individuals. In a SEIR compartmental model individuals are assigned into each group according to their health status: susceptible (S) if they are susceptible to the disease; exposed (E) if they have been infected but are still asymptomatic; infected (I) once the incubation period is over and the individuals show symptoms and can infect others; and removed (R) when they are either recovered or deceased. Within each subpopulation (henceforth, region), the transition between compartments results from the following rules, iterated at each time step, corresponding to 1 day:

**S***→***E:** Susceptible individuals in region *i* might get infected with probability 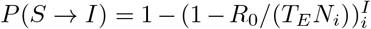, where *R*_0_ is the reproduction number, *T*_*E*_ the mean incubation time, *N*_*i*_ the number of individuals in region *i* and *I*_*i*_ the number of infected individuals in mentioned region.

**E***→***I:** Exposed individuals enter the infected compartment with a rate inversely proportional to the mean latent period,*T*_*E*_.

**I***→***R:** Infected individuals enter the removed compartment with a rate inversely proportional to the mean infectious period, *T*_*I*_.

Note that the generation time is thus *T*_*g*_ = *T*_*E*_ + *T*_*I*_. On the other hand, the mobility of individuals is implemented through a data-driven approach. We have obtained the number of individuals leaving each region each day, 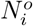, as well as the probability, *p*_*ij*_, of going from each region *i* to region *j* (see Materials and Methods for a thorough description of the data). Hence, at each time step, we select 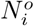 individuals at random from within each region *−* excluding infected individuals in *i*, which are supposed to be under quarantine or hospitalized *−*, and distribute them across the country according to such probabilities. Note, however, that there could be exposed individuals, which are those that ultimately will bring the disease to other subpopulations. We also implemented a randomized version of the model for the mobility, in which the fraction of the population traveling from each region at each time step is 0.008 (estimated from the average fraction of individuals traveling during the first week of January 2020) and their destinations are chosen randomly with probability proportional to the population of the destination region.

Furthermore, since we have data of 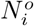 from 2019 and 2020, we simulated the spreading of the disease in both years. In this way, we are able to asses the impact of travel reductions due to mobility restrictions across subpopulations without making any *−* however sensible*−* assumptions about possible changes in individuals’ mobility. Nonetheless, this period of the year has some peculiarities due to the Spring Festival, an event that completely modifies the travel patterns of the population. In 2020, the Spring Festival was celebrated on Jan. 25th, while in 2019 it took place on Feb. 5th. For this reason, we align both simulations so that “day 0” will correspond in both years to the day of the Spring Festival (Jan. 25th and Feb. 5th respectively). As a consequence, we start the simulations on Jan. 12th, 2019 and Jan. 1st, 2020, so that in both years the period between the first reported cases and the celebration of the Spring Festival is the same. Lastly, the simulation will run in both cases to 13 days after the Festival, which corresponds to Feb. 5th in 2020 and Feb. 16th in 2019.

## III. RESULTS AND DISCUSSIONS

Fig. 1 clearly shows that the changes induced by the epidemic in the overall flow and movement of the population throughout China is not restricted just to the region in which the city of Wuhan is located (Hubei). In 2019, a large number of individuals moved right before the Spring Festival. Then, it reached a minimum at the Spring Festival and later on, again, a large number of individuals moved again, see Fig. 1A. On the other hand, in 2020, the situation was similar only until Jan. 23rd, when the travel restrictions in Wuhan were implemented. Two features are worth highlighting. First, when the travel restrictions were implemented, the flow of population had peaked already just before the Spring festival. This is key to understand why, despite the reduction in mobility, the daily number of new cases have continued to increase for weeks. Secondly, most of the second wave observed in 2019 *−* which corresponds to travel back to the original region might have not taken place yet (see Fig. 1B), and thus, there is still a high risk that a subsequent large outbreak or increase in the number of newly infected individuals happens. The existence of recurrent local outbreaks is a feature that some models have anticipated [20]. Further indications that the population has not yet reached the original distribution by region are provided in Figs. 1C and 1D, where we show the number of individuals living in each region at the beginning of the simulation (initial data obtained from the Chinese Yearbook, see Materials and Methods) and at the end according to our data. Clearly, while in 2019 most of the population returned to their regions, in 2020 this has not taken place yet.

**FIG. 1:**
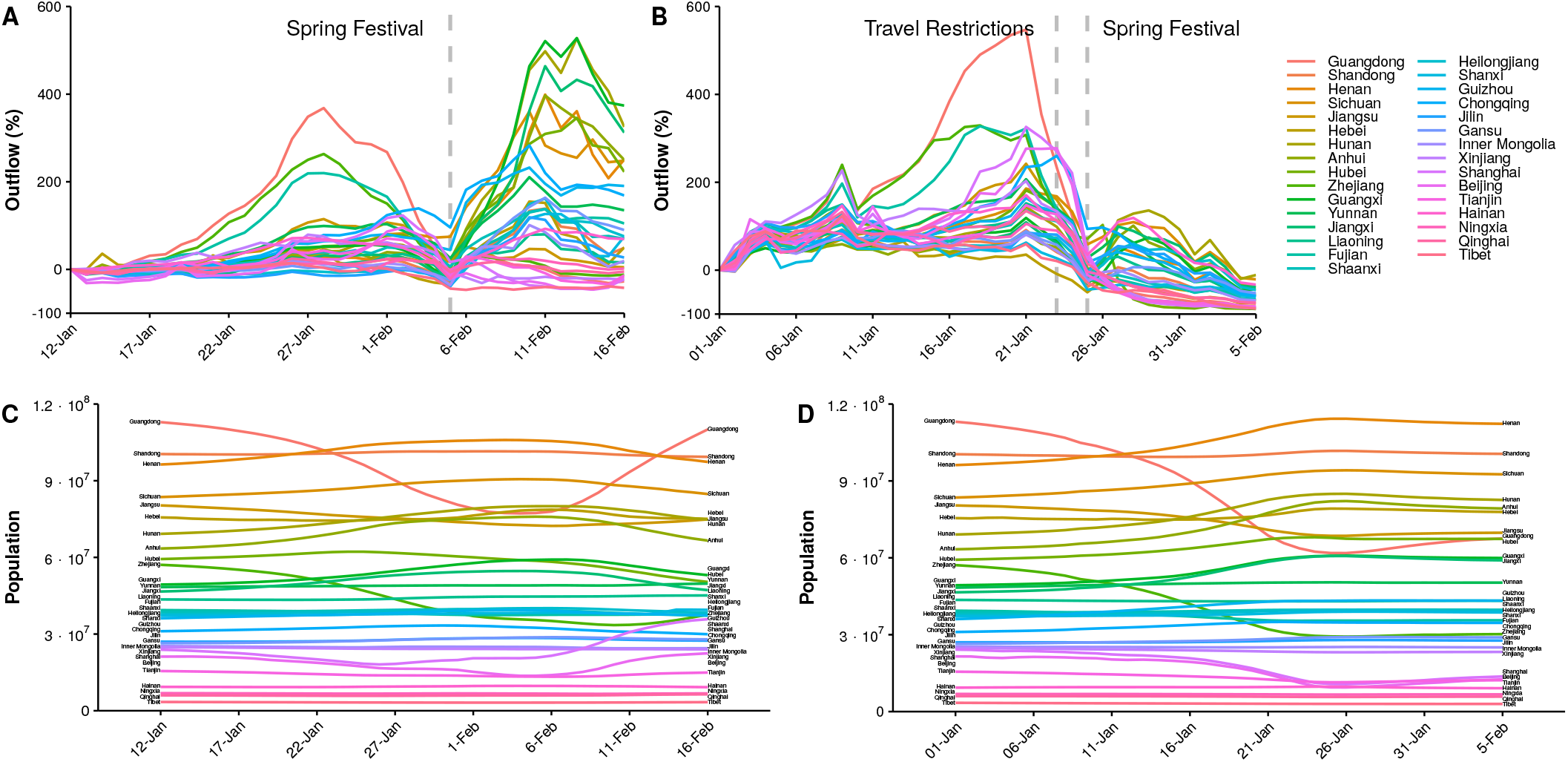
Movement patterns of individuals across Mainland China in 2019 and 2020. Panels A and B represent the percentage of individuals traveling from each region compared to such fraction on the first day of the simulation for 2019 and 2020 (Jan. 12th and Jan. 1st, respectively). Additionally, panels C and D show the population of each region assuming that the number of individuals traveling from each region is given by the flow data and that their destinations are randomly selected according to the mixing patterns *p*_*ij*_.

To parameterize the metapopulation epidemiological model we follow Chinazzi et al. [7] and set a generation time of 7.5 days and a reproduction number *R*_0_ equal to 2.4. The latent period is set to 3 days [21]. Although similar values have been reported by other groups, we have also tested larger values of these parameters and the results are consistent (see Materials and Methods) with those reported in what follows. The outbreak is seeded by introducing 40 exposed individuals on Jan. 1st (Jan. 12th in 2019) [7]. Then, the simulations run for 35 days in both cases and we extract the cumulative number of infected cases in each region as a function of time. Note that as there was no travel restriction in 2019, one can see the results obtained with the 2019 data as the more plausible outcome for the 2020 outbreak should travel restrictions were not adopted. In other words, by comparing 2019 with 2020, we can factor out the impact of the early travel reduction in the city of Wuhan and the subsequent changes in the mobility pattern of the population.

Fig. 2 shows the cumulative number of infected individuals for Hubei and for the rest of mainland China. The large majority of cases, in all situations considered, are contained within Hubei province. However, when we look at the predictions on the rest of the country, we begin to see growing differences between the scenarios analyzed. The results in Fig. 2B also show that travel restrictions have a positive impact in the temporal evolution of the disease (compare 2019 with 2020), in so far the reduction in the flow of individuals delays the spreading of the disease to the rest of mainland China. However, as it can also be deduced from the trend of the curves in Fig. 2B, there is no indication that the growth in the number of cases will evolve following a different functional form. In other words, if no further measures are taken and no modifications are made to the model, one would find that the number of simulated cases at day *τ* in 2019 would be the same Δ days after *τ* in 2020.

**FIG. 2:**
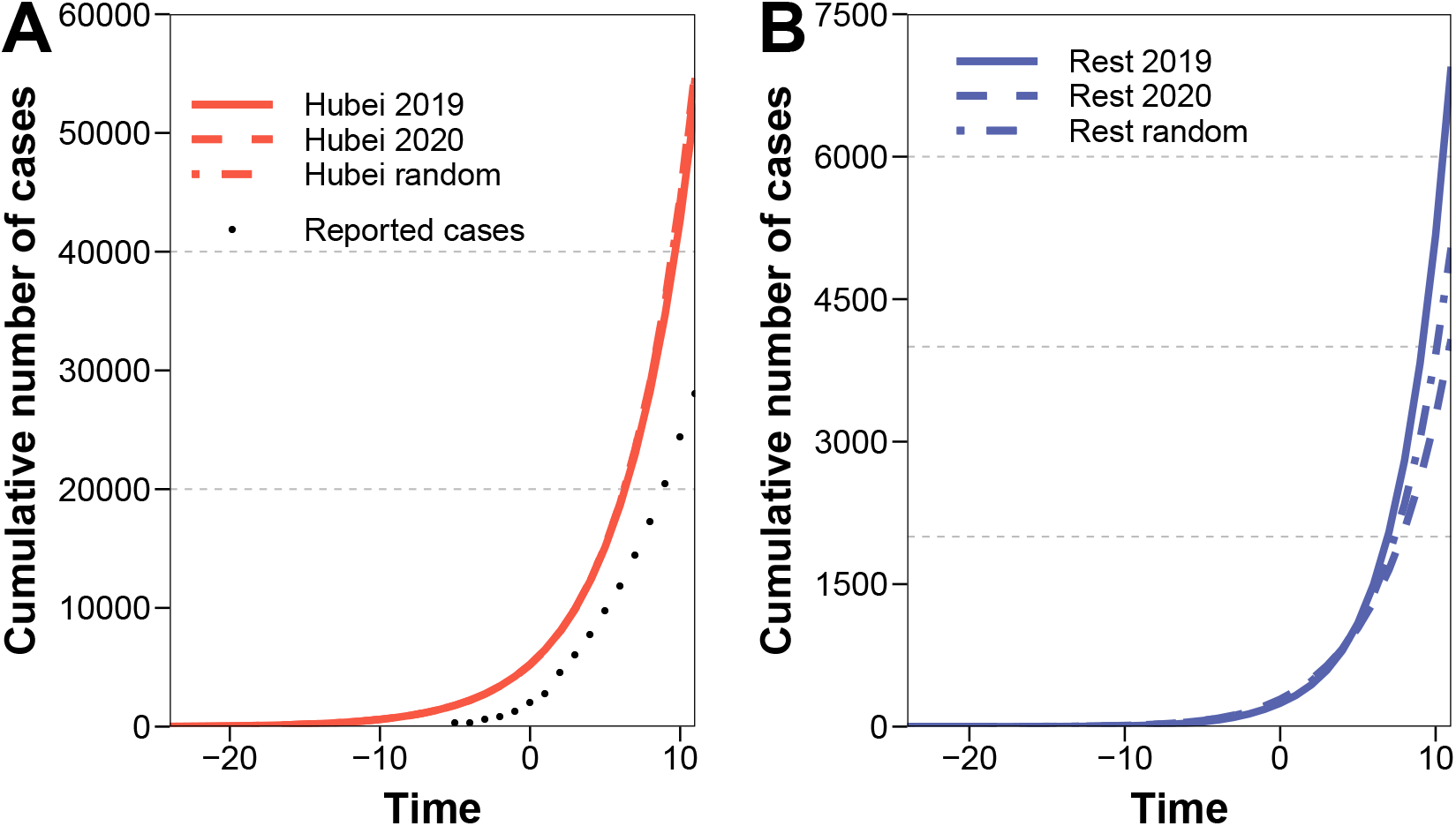
Predicted cumulative number of cases in Hubei (panel A) and the rest of mainland China (panel B) using mobility data of 2019 (solid lines, scenario equivalent to no travel restrictions), of 2020 (dashed lines, with travel restrictions considered) and under the assumption of random movement of the population (dashed-dotted lines). Dots represent the actual total value reported by the authorities.

Fig. 2A also shows that there is a large difference between the cumulative number of cases predicted in 2020 and the reported one. Although, as previously discussed, this has been reported by several other studies, to ensure that the methodology is correct, and to further analyze the effect of the mobility pattern, in Fig. 3 we show the correlation between the real values of infected individuals and the simulated ones for 2020. We obtain a Pearson correlation of 0.81 implying that the assumptions behind the model, albeit simplistic, can correctly describe the basic dynamics of the epidemic. Furthermore, we also see that the data-driven model is able to predict better the dynamics than its random counterpart. Lastly, we have also studied the difference in the predicted number of cases in each region between 2019 and 2020 at the end of the simulation period. Results are reported in Table I, where we show the median as well as the corresponding 5-95% quantiles. As it can be seen in the Table, in the vast majority of the regions the predicted number of cases in the scenario in which travel is not banned (2019) is larger than for those obtained with data from 2020. Interestingly, there are some regions in which the situation reverses. We have no explanation for this (small) negative deviations.

**FIG. 3:**
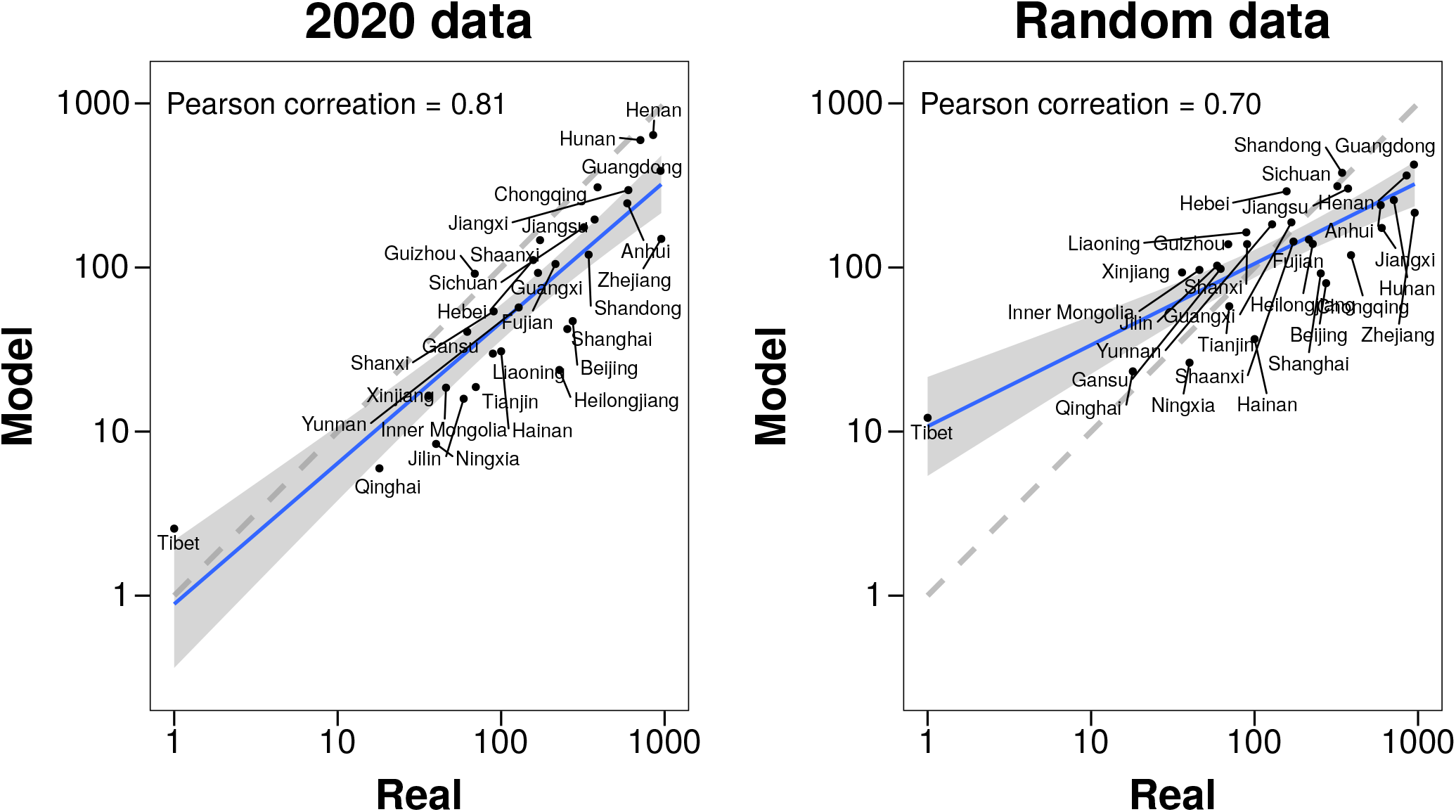
Predicted cumulative number of cases in each region, except Hubei, compared to the real number reported by the authorities by Feb. 5th. Left: estimations obtained using 2020 data. Right: estimations obtained using random mobility data. In both cases the grey dashed line represents the identity line.

**FIG. 4:**
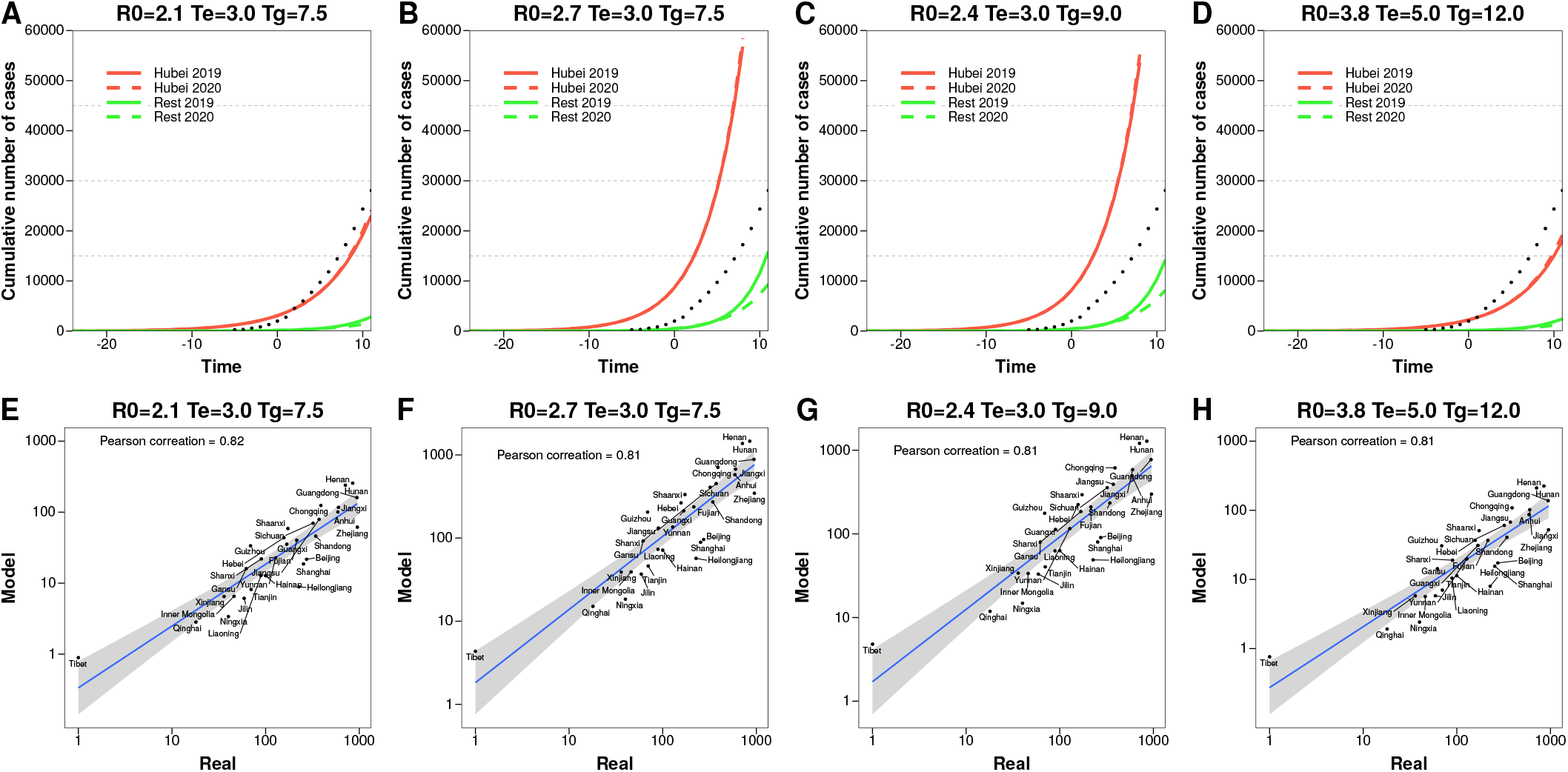
(Top Panels) Predicted cumulative number of cases in Hubei and the rest of mainland China using mobility data of 2019 (solid lines, scenario equivalent to no travel restrictions), and 2020 (dashed lines, with travel restrictions considered) for several parameter values of the epidemic model as indicated. Dots represent the actual total value reported by the authorities. (Bottom Panels) Predicted cumulative number of cases in each region, except Hubei, compared to the real number reported by the authorities by Feb. 5th, 2020 for several disease parameters as indicated.

**TABLE I:** Reported value of the total number of infected individuals by Feb. 5th per region compared to the values predicted by the model using data from 2019 and 2020. The mean value of the prediction, obtained from 104 independent runs of the model, alongside the 5-95% prediction interval is shown. *The outbreak size distribution in Tibet is not well represented by its mean since in most simulations it is 0.

| Region | Reported value | 2019 simulated<br>no travel restriction | 2020 simulated<br>travel restriction |
| --- | --- | --- | --- |
| Henan | 815 | 1,049 [623-1,690] | 642 [231-1,362] |
| Hubei | 19,665 | 52,478 [37,545-68,697] | 55,357 [39,700-72,661] |
| Hunan | 711 | 1,576 [1,022-2,292] | 598 [217-1,254] |
| Guangdong | 944 | 962 [566-1,478] | 388 [156-833] |
| Jiangxi | 600 | 360 [174-701] | 296 [68-744] |
| Chongqing | 389 | 812 [498-1,240] | 308 [84-805] |
| Anhui | 591 | 305 [148-603] | 246 [50-710] |
| Jiangsu | 373 | 198 [66-449] | 196 [36-626] |
| Zhejiang | 954 | 148 [44-350] | 149 [26-456] |
| Shaanxi | 173 | 285 [146-514] | 147 [18-483] |
| Sichuan | 321 | 307 [159-562] | 176 [30-554] |
| Beijing | 274 | 79 [31-160] | 47 [5-57] |
| Shanghai | 254 | 129 [57-254] | 42 [5-54] |
| Shandong | 343 | 90 [24-262] | 120 [8-402] |
| Fujian | 215 | 87 [19-252] | 105 [8-384] |
| Guizhou | 69 | 57 [11-177] | 92 [2-352] |
| Hebei | 157 | 82 [21-233] | 111 [5-413] |
| Guangxi | 168 | 77 [19-212] | 93 [4-335] |
| Yunnan | 128 | 46 [8-147] | 57 [1-253] |
| Shanxi | 90 | 124 [72-207] | 54 [0-236] |
| Hainan | 100 | 16 [0-65] | 31 [0-131] |
| Liaoning | 89 | 25 [4-72] | 30 [0-134] |
| Gansu | 62 | 29 [4-91] | 41 [0-188] |
| Tianjin | 70 | 26 [5-77] | 19 [0-90] |
| Heilongjiang | 227 | 15 [1-49] | 24 [0-110] |
| Xinjiang | 36 | 12 [0-46] | 17 [0-93] |
| Inner Mongolia | 46 | 18 [2-48] | 19 [0-77] |
| Jilin | 59 | 20 [2-56] | 16 [0-74] |
| Qinghai | 18 | 5 [0-17] | 6 [0-22] |
| Ningxia | 40 | 6 [0-23] | 8 [0-36] |
| Tibet | 1 | 1 [0-4] | 3 [0-1]* |

Summarizing, we have studied a data-driven metapopulation model that allows assessing the effects of early travel restrictions in Wuhan and Hubei province. Even if our modeling framework is simpler than other more sophisticated implementations of the disease dynamics, our results are in line with several available studies in that: *i)* travel restrictions are efficacious in the short term, effectively decreasing the expected number of infected individuals in most of the regions in mainland China, and *ii)* at the same time, reducing the travel does not appear to have a long term impact on the spreading of the disease if not accompanied by other measures. We also note that the observed shift of the epidemic curve and the effect of travel restriction might be underestimated because the large majority of people had already moved before these mobility restrictions were implemented (as it can be seen in Fig. 1). Our study is limited in several aspects that can constitute future research goals. First, the geographic resolution allowed by the mobility data used here is low. Considering large regions have the undesired effect that one can not add structure to the population and therefore the dynamics within each subpopulation is constrained by the homogeneous mixing hypothesis. This limitation could be overcome if less granular spatial and temporal data becomes available. Secondly, and perhaps more important as it currently represents a scientific challenge, we have assumed that the transmissibility does not change during the whole simulation period. This implies that changes in behavioral patterns of the population are not fully accounted for nor they can be completely disentangled from those associated with travel restrictions. Understanding how to deal with such behavioral changes is key for the development of more realistic descriptions of the large-scale spreading of diseases. Finally, another critical feature of current models that needs to be improved in future research is the use of disease parameters notably *R*_0_ that are constant both in time and across populations.

## Data Availability

All the data used in this article can be downloaded from their original source. The links can be found in the references list.

## Acknowledgments

YM acknowledges partial support from the Government of Aragon, Spain through grant E36-17R (FENOL), and by MINECO and FEDER funds (FIS2017-87519-P). AA and YM acknowledge support from Intesa Sanpaolo Innovation Center. AA and QH share cofirst authorship. PJ acknowledges Natural Science Foundation of Shanghai, Eastern Scholar and NSFC 269 (11701096). The funders had no role in study design, data collection, and analysis, decision to publish, or preparation of the manuscript.

## MATERIALS AND METHODS

### Data description

#### Resident population

The statistics about the distribution of the population across 31 regions in China were obtained from the “China Statistical Yearbook 2019” [22]. This annual publication reflects comprehensively the economic and social development of China.

#### Population flow

We obtained migration data from Baidu Quanxi, an open platform based on Baidu Location Based Services (LBS) that provides information about the population flow within China [23]. The dataset comprises two types of data: the Baidu migration index and the Baidu migration ratio. The former is a number proportional to the number of individuals leaving each region. To obtain the proportionality constant, we averaged the amount of individuals leaving Wuhan between Jan. 1st and 10th and compared it to the value of 502,013 estimated by Wu et al. [15]. The second part of the dataset contains the fraction of individuals going from region *i* to region *j, p*_*ij*_. As such, it is possible to estimate the number of individuals going from region *i* to *j* by multiplying the total outflow of the region by the corresponding *p*_*ij*_. The outflow of the individuals is available for the years 2019 and 2020, while the *p*_*ij*_ values only for 2020. Nevertheless, as it can be seen in Fig.1 in the main text, it represents a good proxy of the situation in 2019. Indeed, even though some populations were closed after Jan. 23rd, 2020, using these values for 2019 correctly describes the return of the population that had left for the Spring Festival.

#### Infected individuals

The number of infected individuals as a function of time, as well as their distribution across regions on Feb. 5th was obtained from the WHO reports [1].

### Sensitivity analysis

To gauge the effect of the chosen parameterization of the model, we have repeated the analysis with several values reported in the literature [7, 24]. As expected, if the incubation period is larger or the basic reproduction number is smaller, the overall number of cases decreases. Conversely, when using large values of *R*_0_ or longer generation times, the amount of infected individuals increases. In any case, the mobility model is quite independent from the dynamics of the disease, since the correlation is almost always the same in all cases considered.

## Notes

### Competing Interest Statement

The authors have declared no competing interest.

## References

[1] Tech. Rep., World Health Organization (2020), URL https://www.who.int/docs/default-source/coronaviruse/situation-reports/20200216-sitrep-27-covid-19.pdf.

[2] G. S. Randhawa, M. P. M. Soltysiak, H. E. Roz, C. P. E. de Souza, K. A. Hill, and L. Kari, bioRxiv p. 2020.02.03.932350 (2020), 2020.02.03.932350.

[3] L. Wahba, N. Jain, A. Z. Fire, M. J. Shoura, K. L. Artiles, M. J. McCoy, and D. E. Jeong, bioRxiv p. 2020.02.08.939660 (2020), 2020.02.08.939660.

[4] P. Zhou, X.-L. Yang, X.-G. Wang, B. Hu, L. Zhang, W. Zhang, H.-R. Si, Y. Zhu, B. Li, C.-L. Huang, et al., Nature pp. 1–4 (2020), ISSN 1476-4687.

[5] K. Gostic, A. C. R. Gomez, R. O. Mummah, A. J. Kucharski, and J. O. Lloyd-Smith, eLife (2020).

[6] Z. Du, L. Wang, S. Cauchemez, X. Xu, X. Wang, B. J. Cowling, and L. A. Meyers, medRxiv p. 2020.01.28.20019299 (2020).

[7] M. Chinazzi, J. T. Davis, M. Ajelli, C. Gioannini, M. Litvinova, S. Merler, A. P. y. Piontti, L. Rossi, K. Sun, C. Viboud, et al., medRxiv p. 2020.02.09.20021261 (2020).

[8] Z. Cao, Q. Zhang, X. Lu, D. Pfeiffer, L. Wang, H. Song, T. Pei, Z. Jia, and D. D. Zeng, medRxiv p. 2020.02.07.20021071 (2020).

[9] Y. Zhou and J. Dong, medRxiv p. 2020.02.10.20021774 (2020).

[10] X. Zhu, A. Zhang, S. Xu, P. Jia, X. Tan, J. Tian, T. Wei, Z. Quan, and J. Yu, medRxiv p. 2020.02.09.20021360 (2020).

[11] Tech. Rep., World Health Organization (2020), URL https://www.who.int/docs/default-source/coronaviruse/situation-reports/20200213-sitrep-24-covid-19.pdf.

[12] Tech. Rep., World Health Organization (2020), URL https://www.who.int/docs/default-source/coronaviruse/situation-reports/20200212-sitrep-23-ncov.pdf.

[13] X. Li, X. Zhao, and Y. Sun, medRxiv p. 2020.02.09.20021477 (2020).

[14] H. Xiong and H. Yan, medRxiv p. 2020.02.10.20021519 (2020).

[15] J. T. Wu, K. Leung, and G. M. Leung, Lancet 0 (2020), ISSN 0140-6736.

[16] H. Tian, Y. Li, Y. Liu, M. U. G. Kraemer, B. Chen, J. Cai, B. Li, B. Xu, Q. Yang, P. Yang, et al., medRxiv p. 2020.01.30.20019844 (2020).

[17] H.-Y. Yuan, M. P. Hossain, M. M. Tsegaye, X. Zhu, P. Jia, T.-H. Wen, and D. Pfeiffer, medRxiv p. 2020.02.01.20019984 (2020).

[18] P. Boldog, T. Tekeli, Z. Vizi, A. Denes, F. Bartha, and G. Rost, medRxiv p. 2020.02.04.20020503 (2020).

[19] V. Colizza and A. Vespignani, J. Theor. Biol. 251, 450 (2008), ISSN 0022-5193.

[20] A. Aleta, A. N. S. Hisi, S. Meloni, C. Poletto, V. Colizza, and Y. Moreno, R. Soc. Open Sci. (2017).

[21] W.-j. Guan, Z.-y. Ni, Y. Hu, W.-h. Liang, C.-q. Ou, J.-x. He, L. Liu, H. Shan, C.-l. Lei, D. S. Hui, et al., medRxiv p. 2020.02.06.20020974 (2020).

[22] CSYD, China Statistical Yearbook (accessed February 24, 2020), http://www.stats.gov.cn/tjsj/ndsj/2019/indexeh.htm.

[23] Baidu, Baidu Qianxi (accessed February 24, 2020), qianxi.baidu.com.

[24] Y. Yang, Q. Lu, M. Liu, Y. Wang, A. Zhang, N. Jalali, N. Dean, I. Longini, M. E. Halloran, B. Xu, et al., medRxiv p. 2020.02.10.20021675 (2020).

